# Awareness and indicators of low energy availability in male and female dancers

**DOI:** 10.1101/2020.06.28.20141580

**Authors:** Nicola Keay, AusDancers Overseas, Gavin Francis

## Abstract

**Objectives:** To investigate awareness and indicators of low energy availability (LEA) in male and female dancers

**Methods:** A dance-specific energy availability questionnaire (DEAQ) was developed and administered online internationally to dancers in full time training. The DEAQ drew on current validated, published questionnaires for LEA, linked to the clinical outcomes of relative energy deficiency in sport (RED-S). Questions addressed recognised physiological indicators and consequences of LEA in the context of dance, together with psychological drivers and aetiological factors specific to dance training. LEA was quantified using a scoring system to include these characteristics.

**Results:** 247 responses to the DEAQ were analysed (225 female and 22 male), mean age 20.7 years (SD 7.9) with transition to full time training at 15.0 years (SD 7.9) and 85% practising ballet. Psychological, physiological and physical characteristics consistent with LEA were reported by 57% of the female dancers and 29% of male dancers, indicating a risk of RED-S. The unique nature of dance training, in terms of demands and environment, was found to be potentially influential in development of this situation. Less than a third (29%) of dancers were aware of RED-S.

**Conclusion:** This study found dancers to be a specific group of high-level exercisers displaying indicators of LEA and consequently at risk of developing the adverse clinical health and performance consequences of RED-S. Awareness of RED-S was low. The DEAQ has the potential to raise awareness and be a practical, objective screening tool to identify dancers in LEA, at risk of developing RED-S.

**Summary boxes:** *What are the new findings?:*

- Dancers reported many recognised indicators of low energy availability (LEA) and consequently are at risk of developing the adverse health and performance outcomes of relative energy deficiency in sport (RED-S).
- Few dancers in this study demonstrated an awareness of RED-S.
- The unique nature of dance training, in terms of demands and environment, was found to be influential in the development of LEA in dancers
- The DEAQ is the first questionnaire specific to dancers. Applying a scoring system to the responses from the DEAQ can provide an objective assessment of LEA

*How might this study impact on clinical practice in the future?:*

- As LEA and subsequent risk of RED-S is not matched by awareness, providing educational resources for dancers is important. A British Association of Sport and Exercise Medicine website has been developed by the research team for both athletes and dancers www.health4performance.co.uk
- The DEAQ has the potential to be a practical, objective, screening tool to identify male and female dancers worldwide in LEA. By identifying these dancers, support could be targeted to modify dancer behaviours to reduce the risk of dancers developing the adverse health and performance sequelae of RED-S
- Early identification of dancers at risk of developing RED-S is of particular importance when situations arise out of dancers’ control, such as lock down in pandemics COVID-19 or time off dancing due to illness/injury. Targeted support may be required as a dancer’s tendencies towards exercise dependence and disordered eating patterns may increase as a way to seek control and combat uncertainty.

## Introduction

Low energy availability (LEA) causes physiological and psychological disruption leading to adverse health and performance outcomes described in the clinical model of relative energy deficiency in sport (RED-S) [1,2]. RED-S is prevalent in sports where low body weight confers a performance or aesthetic advantage[2].

Direct measurement of energy availability (EA) is not practical as a screening tool being laborious, valid only at the time of assessment and prone to inaccuracies. Furthermore, threshold EA is not universally applicable for individuals[3]. Physiological indicators of LEA are clinically relevant and assessed in a validated questionnaire for female athletes: LEAF-Q[4]. LEA measured using a self-reported questionnaire is strongly associated with many health and performance consequences of RED-S[32]. However, these questionnaires exclude males and underlying psychological drivers for LEA, nor are they exercise-specific. In male athletes, a self-reported questionnaire found evidence of exercise hypogonadal condition and other physiological indicators of RED-S[35]. A sport-specific energy availability questionnaire combined with interview (SEAQ-I)[5] for male cyclists was effective in indicating low bone mineral density (BMD) of the lumbar spine, characteristic of RED-S.

Endocrine disruption, menstrual status in women and testosterone levels in men, indicate LEA, which is linked to the clinical outcome in RED-S of impaired bone health and stress fracture in runners[6]. The IOC RED-S Clinical Assessment Tool (RED-S CAT)[7] is applicable to male and female athletes, but is designed for medical professional use only. Combining these approaches could be of great clinical value for dancers, who share the physical demands of athletes and for whom low body weight is considered a performance and aesthetic requirement. Dancers are potentially at risk of RED-S, yet the majority of research to date has focused on sports. The objectives of this study were to establish the prevalence of indicators of LEA in dancers, in order to assess contributing factors, awareness and risk of RED-S in dance.

## Methods

### Study design

A cross sectional, observational study of a newly developed questionnaire was designed to assess the risk of RED-S in male and female dancers. The study was approved by the university ethics committee and participants provided informed consent before completing the questionnaire.

### Recruitment

Participants were recruited to the study through contacts of the researchers with dancers and dance organisations. Dancers were eligible to take part in the study if trained and performed at pre-professional, professional or advanced amateur level.

### Dance-specific energy availability questionnaire (DEAQ)

The DEAQ drew upon validated questionnaires, including LEAF-Q[4], SEAQ-I[5], androgen deficiency[8] and RED-S CAT[7]. Dancer descriptive characteristics included age and self-reported body weight and height. Questions covering training, attitudes to weight and eating behaviours were tailored to be dance specific. The presentation and order of questions were considered to ensure engagement from dancers. The questionnaire was reviewed by the Sports Sciences Department at Durham University, including a psychologist and dance medicine endocrinologist; medical and nutritional professionals at AusDancers Overseas and dancers for content validity.

The DEAQ (supplementary file 1) was administered online using the Jisc survey tool. A RED-S Risk Score was formulated, applying a points system to DEAQ responses particularly indicative of LEA, as shown in supplementary file 2. Relevant factors included physical indicators, such as body mass index (BMI), lowest weight, injuries; physiological factors, including indicators of sex steroid hormone levels, gut issues; psychological aspects including dietary and exercise behaviours, measures of wellbeing (freshness, sleep), attitudes to controlling exercise, diet and weight and any history of a diagnosed eating disorder.

### Statistical analysis

Data analyses were performed using the open source tool, Pandas (NumFOCUS, Austin, Texas), implemented using the Python programming language. Summary statistics, including count, mean and standard deviation of responses, were calculated for the overall sample and by subgroup, according to the responses to the questions.

Body mass index (BMI) was calculated by dividing weight (kg) by the square of height (m). Minimum BMI (BMI min) was calculated based on the dancer’s minimum weight for current height. A weight variability variable was calculated for the current height by dividing the difference between the maximum and minimum weights by the current weight.

### Public and patient involvement

Public involvement was integral at all stages of this research. The conception of an objective survey arose from discussions with dancers. Whilst the questions were based on validated questionnaires, dancers contributed to drafting questions in the context of dance training and suggesting extra dance specific questions. Anonymisation of responses was agreed. Dancer networks supported recruitment and will support dissemination of the findings with independent dance organisations, dance publications and international dance meetings.

## Results

### Participant characteristics

The questionnaire received responses from 247 dancers (225 women and 22 men) The mean age of women was 20.5 years (SD 8.0) and men 22.5 years (SD 7.1). On average, dancers started aged 5.8 (SD 4.1) and transitioned to full-time training at 15 years (SD 3.1). Ballet was the main form of dance in 85% of female and 91% of male dancers. Table 1 shows reported anthropomorphic data. Weight variability of 15% would be consistent with a dancer weighing 53.9kg seeing her adult weight fluctuate between 50kg and 58kg and her BMI range from 18.2 to 21.2 kg/m^2^.

**Table 1:** Anthropomorphic data of dancers.

| Mean and range | Height (m) | Weight (kg) | BMI (kg/m2) | Weight variability* |
| --- | --- | --- | --- | --- |
| **Women (n=225)** | 1.66 [1.47 to 1.83] | 53.9 [29.5 to 86.7] | 19.7 [13.1 to 31.1] | 15% [0% to 53%] |
| **Men (n=22)** | 1.77 [1.57 to 1.87] | 67.3 [42.0 to 82.0] | 21.5 [17.0 to 25.3] | 9% [0% to 32%] |

Table footnote: \*weight variability calculated for current height by dividing the difference between maximum and minimum weight by current weight

A quarter of dancers had been engaged in on average 10.4 hours (SD 7.1) a week sports training prior to taking up full time dancing. Table 2 shows the current weekly activities of the dancers.

**Table 2:** Current weekly activity levels.

| **Hours per week**<br>**Mean and range** | **Class** | **Rehearsal** | **Performance** | **Conditioning/Gym**<br>**work/Supplementary**<br>**training** | **Total** |
| --- | --- | --- | --- | --- | --- |
| **Women** | 15.3 [0 to 47] | 9.8 [0 to 43] | 1.9 [0 to 16] | 5.1 [0 to 35] | 29.9 [0 to 80] |
| **Men** | 15.0 [2 to 40] | 12.0 [0 to 37] | 2.3 [0 to 9] | 3.2 [0 to 8] | 30.4 [0 to 50] |

### Psychological factors

#### Attitudes to training

71% of respondents reported feeling worried about missing a session, with common reasons being missing out on learning choreography and potential roles, loss of technique, guilt or absence noticed by teachers.

#### Attitudes to weight

44% of women and 32% of men reported being told to lose weight at some point in their training/professional career, mainly by teaching staff/directors. 83% of dancers were influenced to some extent by social media in trying to lose weight, with 30% of women and 14% of men saying this was a constant influence.

Dancers reported weighing themselves on average 1.8 times a week. 42% did not track their weight, while fewer than 10% weighed themselves more than 3 times a week. Using a scale of 1 (no effect) to 6 (very important), female dancers felt that their ability to control what they eat (4.6) and what they weigh (4.1) affected their self-esteem more than men, 4.0 and 3.1, respectively.

Dancers generally considered their best dance weight to be lower than their current weight: on average, women would prefer to lose 1.8kg and men 1.3kg. 71% of women and 43% of men agreed that a leading role would favour someone of lower weight for height.

### Endocrine status

Male hormone status was assessed by the number of morning erections, where the average was 4.2. Female hormone history was investigated in greater detail (Figure 1).

**Figure 1.**
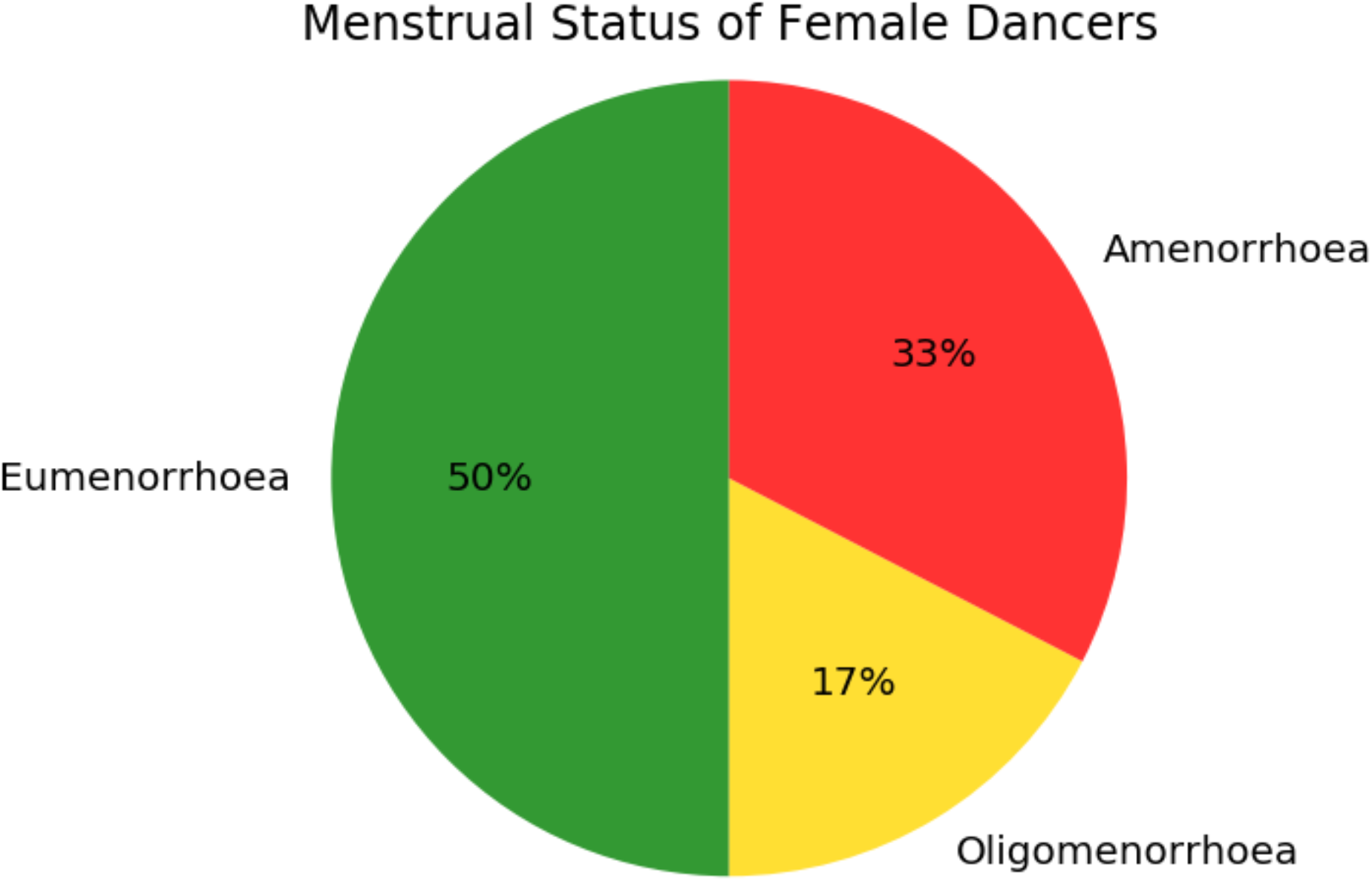
Menstrual Status in Female Dancers

Primary amenorrhoea: 8% of women reported that menstruation had never started. For 19%, periods did not begin until age 15 or later. Of 184 women not taking hormonal contraception, only half reported having 9 or more cycles per calendar year, while one third experienced primary (n=14) or secondary amenorrhoea (n=46), with the remaining group having oligomenorrhoea (n=32). 28% had experienced three or more consecutive months without periods (besides pregnancy or taking hormonal contraception) and a further 26% stated that this was the current situation. 58 women had reported to school/company a lack of periods, in 43% of cases (n=25) the matter was not addressed.

15% of women (n=34) were taking hormonal contraceptives. Of these, half were seeking to prevent pregnancy. Other reasons included reducing bleeding (35%), regulating cycles in relation to performances (32%), reducing menstrual pain (14%), to induce monthly bleeds (12%) or medical reasons such as endometriosis (18%).

90% of women understood that that the hormones in the oral contraceptive pill are not equivalent to the body’s hormones. 57% of male and female participants thought it abnormal for female dancers not to have periods, 23% thought it normal and 19% did not know. 79% of women acknowledged negative consequences from not having periods (apart from not being able to get pregnant), 6% disagreed and 16% did not know; while 68% of men agreed and 32% did not know.

### Illness and injuries

Most dancers missed relatively few days due to illness or injury, but a small minority had suffered recurrent issues (Table 3). Soft tissue injuries were more prevalent than bone injuries, with a higher incidence of recurrence. Bone fractures were most common in lower limb, pelvis and spine. Among females who had experienced an injury, healthcare professionals enquired about periods in 35% of cases.

**Table 3:** Illness and injuries in dancers.

| **Mean and range** | **Days off dancing due to illness in last year** | **Days off dancing due to injury in last year** | **Soft tissue injuries from dancing** | **Recurrent soft tissue injuries** | **Bone injuries from dancing** | **Recurrent bone injuries** |
| --- | --- | --- | --- | --- | --- | --- |
| **Women** | 6.3 [0 to 180] | 13 [0 to 200] | 1.5 [0 to 14] | 1.0 [0 to 7] | 0.6 [0 to 6] | 0.4 [0 to 7] |
| **Men** | 3.5 [0 to 20] | 10 [0 to 80] | 2.1 [0 to 10] | 1.5 [0 to 5] | 0.5 [0 to 2] | 0.3 [0 to 2] |

### Eating habits

23% of women and 5% of men were vegetarian, while 10% of male and female dancers were vegan. 53% of women and 36% of men excluded certain foods from their diets, most commonly meat, followed by carbohydrates. 98 dancers had been advised by teachers/dancers to exclude foods, most frequently carbohydrates.

Popular sources for nutrition information were the Internet (61%), professional dietician/nutritionist (32%), friends/teachers at school/company (29%), school/company provided access to dietician/nutritionist (17%). 11% did not seek advice on nutrition.

#### Supplements, allergies/intolerances

77% of dancers reported taking supplements, most commonly vitamins D and B12. 80 dancers reported allergies/intolerances, 33% of women and 27% of men, with 28 reporting being tested.

### Wellbeing

On a rating scale for freshness from 1 (extremely fatigued) to 6 (no fatigue at all), on average women scored 3.7 and men 3.8. For sleep, where 1 (hardly ever get a good night’s sleep) to 6 (always), men and women both ranked 4.0 on average. Among those having problems sleeping, common reasons were difficulty falling asleep (37%), disrupted sleep (21%) and early waking (18%).

Ratings for the digestive system, from 1 (continuous problems) to 6 (none), produced an average for women of 4.2 and men 4.5. 23% reported no digestive problems, while 35% had bloating, 23% suffered discomfort and 12% constipation.

### Eating disorders

Eating disorders had been diagnosed in 15% of women and 14% of men. These included anorexia nervosa (64%), bulimia (22%). Disordered eating: orthorexia (8%).

### Awareness

29% had heard of RED-S and 37% low energy availability. 30% had come across the female athlete triad. 90% were aware of terms disordered eating, 46% orthorexia.

### RED-S Risk Scores

RED-S risk scores ranged from −17 to +16, with negative scores indicating lower energy availability and therefore higher risk of RED-S. The average score for women (Figure 2) was −1.4 (SD 6.1) and +2.3 (SD 6.0) for men (Figure 3). Negative scores were seen in 57% female dancers and 29% male dancers. Women with the modal score of −5 were characterised as having relatively low BMI (19.0) and BMI min (17.3), often had hormone issues (late menarche, irregular or disrupted cycles), over a week off due to injury and strong desires to control diet (5.2 out of 6) and weight (4.6 out of 6).

**Figure 2.**
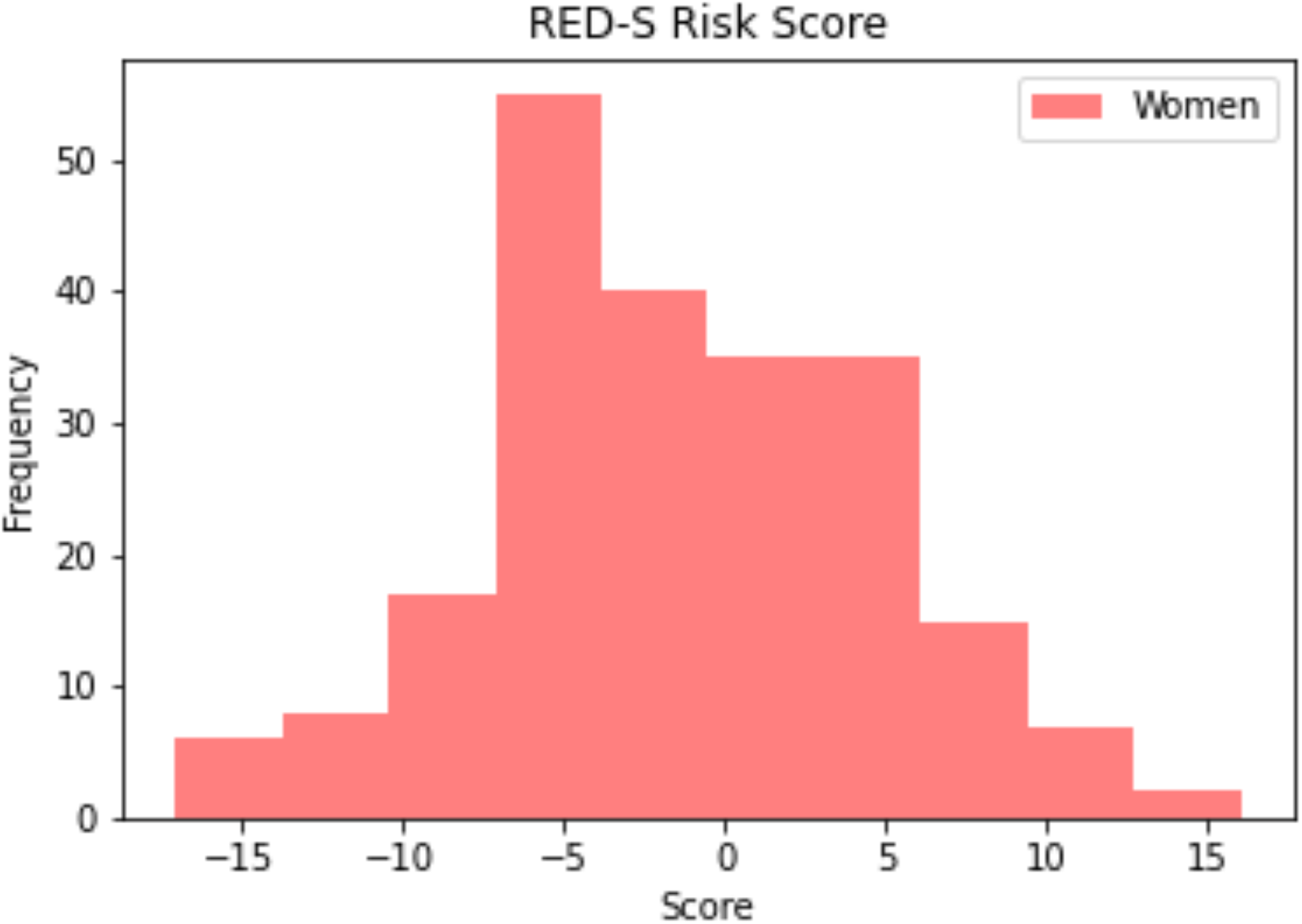
RED-S risk score for Female Dancers

**Figure 3.**
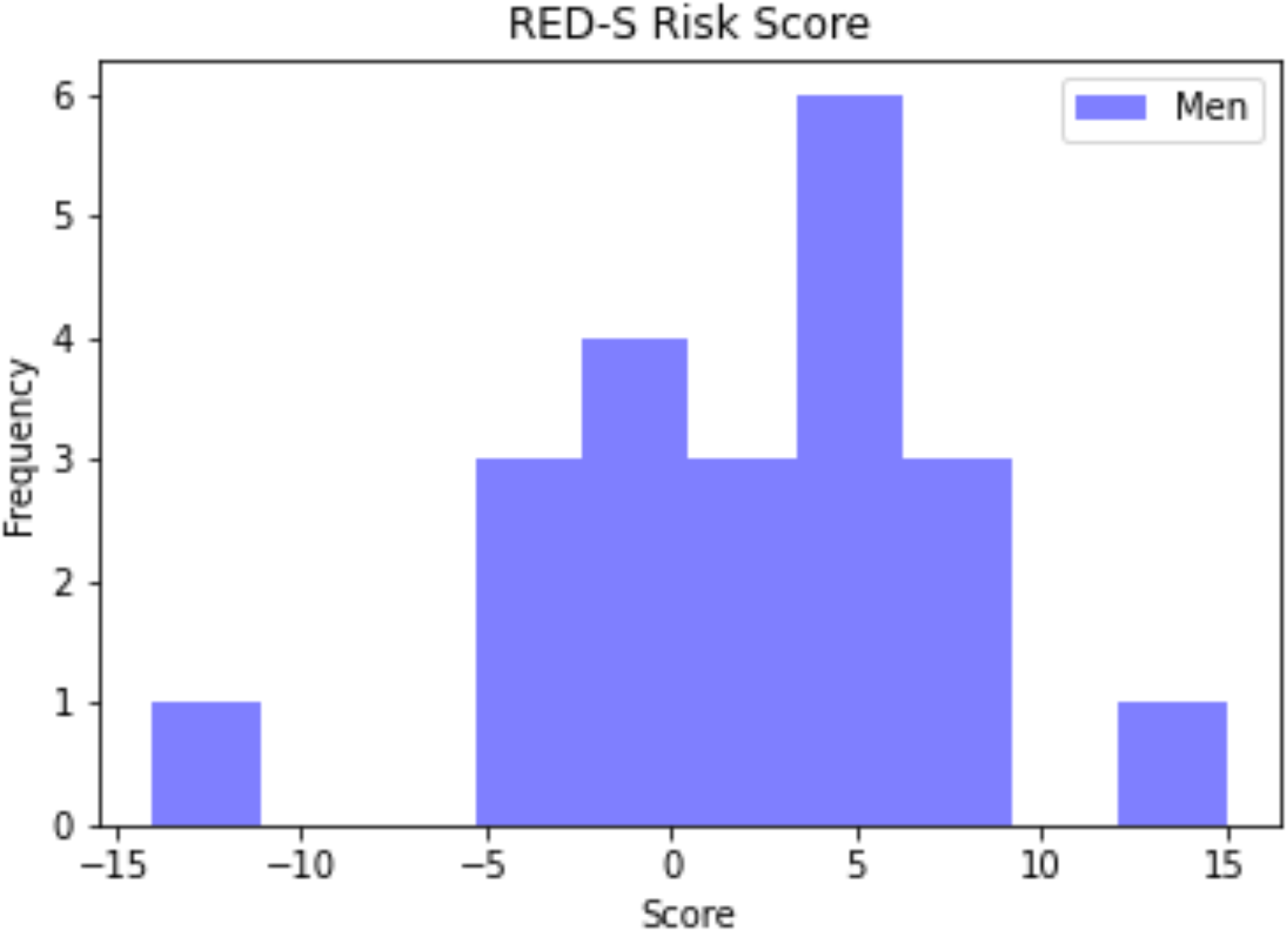
RED-S risk score for Male Dancers

## Discussion

A dance-specific questionnaire (DEAQ) investigating physical, physiological and psychological characteristics showed that dancers are a specific group of high-level exercisers displaying indicators of LEA.

### Physical aspects

Ballet, the main dance form in this study, is practised worldwide and requires a slim physique to meet aesthetic and technical demands. Early training specialisation (full time training commencing, on average, aged 15) combined with a high training load (average of 29.9 hours per week), found in this study, is characteristic of dance[9]. Intensive training during the teenage years increases the already considerable energy demands of physical development. This is recognised as an early risk factor for developing RED-S[10, 11]. Our study found BMI at the lower end of range (average for female dancers 19.7) and large variation in adult weight (15% for female dancers). In a study of retired female dancers, low BMI and duration of amenorrhoea during a dance career were found to be independent variables for low bone mineral density in the long-term[12].

### Psychological factors

Psychological factors can both contribute to the cause and be the consequence of LEA[1,2]. A detrimental interplay can be perpetuated by the nature of dance training, where selection strongly favours certain physical and psychological characteristics.

Psychological drivers of competitiveness, perfectionism and self-control can render athletes susceptible to disordered eating behaviours[13]. Development of attitudes regarding “ideal” body type/weight and teammate modelling of eating behaviours can trigger disordered eating in junior athletes and pre-professional dancers[14, 15]. Vocational training involves living away from home, often abroad, where dancers may use dance peers and social media as comparators.

The drive for thinness is an indicator of LEA, with increased menstrual disruption[16], reduced triiodothyronine and resting metabolic rate(RMR)[17]. Reduced RMR in male and female dancers is an indicator of LEA[18]. Dancers in this study expressed dissatisfaction with their current weight, wishing to be lighter.

The perceived performance advantage of weight loss can be a driver of disordered eating in aesthetic sports and dance [19, 20]. In our study, 73% of female dancers stated that being of low body weight would improve chances of being cast in significant roles. Both male and female dancers indicated that controlling what they ate and what they weighed were important factors linked with self-esteem.

44% of female dancers and 33% of males reported being advised, at some point, to lose weight. Most commonly, dancers had been encouraged to exclude carbohydrates, contrary to research showing that low carbohydrate diets limit physical performance at high intensities[21].

83% of dancers cited social media as influential in feeling that weight loss was desirable. “Thinness related learning” for dancers, who already have perfectionist traits, is cited as increasing the risk for eating disorders[22]. This study revealed a high incidence of a previous eating disorder, 15 % for female and 14% for male dancers, primarily anorexia nervosa. These findings are comparable to those previously reported in dancers and athletes, being higher than the non-athletic population[23, 24]. Exclusion of food groups by choice was reported by 50% female and 33% of male dancers

Exercise dependence is reported as a reliable indicator of eating psychopathology tendencies in female athletes[25] and biochemical indicators of RED-S in male athletes[26]. 71% of dancers in this study reported feeling anxious about missing class.

Once athletes fall into LEA accompanied by resultant disrupted hormone networks, the psychological consequences reinforce and perpetuate these problematic belief systems and behaviours[2].

### Physiological outcomes

#### Endocrine function

Menstrual status in women and testosterone levels in men, are sensitive, objective indicators of LEA, linked to the clinical outcome of impaired bone health and stress fracture of RED-S[6]. Menstrual disruption, in particular functional hypothalamic amenorrhoea due to LEA, is characteristic of the clinical risk assessment of RED-S[7]. The high incidence of menstrual disruption amongst dancers in this study, is far in excess of general population[27]: primary amenorrhoea 8% versus less than 0.01% and combined primary and secondary amenorrhoea 33% versus 4%. A past history of amenorrhoea was reported 28% in dancers. Oligomenorrhoea associated with increase in training loads was reported in 17% of dancers. Overall, half of the female dancers reported disrupted menstrual function. Extensive literature demonstrates adverse health outcomes of the hypo-oestrogen state in terms of impaired bone, cardiovascular and neuromuscular function[28, 29, 30]. Although 79% female dancers recognised that lack of menstrual cycles could have adverse consequences; 23% considered this “normal” for dancers. This erroneous view might be perpetuated by a failure of staff to address amenorrhea (43%) or enquire about menstruation on presentation of injury (35%), despite amenorrhoeic status being a recognised risk factor for both soft tissue and bone stress injuries[6].

#### Gastrointestinal function

Disrupted gastrointestinal function is a well documented, validated indicator of LEA[4]. 77% dancers reported such digestive issues indicative of LEA. Misinterpretation of these symptoms can prompt further restrictive practices: half of female dancers and a third of male dancers were excluding food groups. Furthermore a third of dancers reported food intolerances, although very few had actually been formally tested.

#### Injuries

Incidence of injury was lower than might be expected, in view of the high percentage of dancers assessed as being in LEA. The young average age of participants might be a factor, as the consequences of LEA occurring during peak bone mass accumulation manifest with an increased incidence of stress fractures during mid-20s[31]. Furthermore, participants in this study were not pre-selected by attending an injury clinic where higher incidences were reported[32].

### RED-S risk score from DEAQ

Negative scores from the DEAQ indicated LEA in 57% of female and 29% of male dancers, suggesting that these dancers are at risk of developing the clinical consequences of RED-S. For females athletes LEA assessed by questionnaire is well documented as a validated tool to indicate risk of developing the adverse clinical outcomes of the female triad (poor bone health, menstrual disruption and disordered eating)[4] and RED-S, where in addition to triad symptoms, includes negative multisystem and performance consequences[32]. Comparison of LEA risk quantification in female athletes using RED-S CATs[7] and Female Athlete Triad Cumulative Risk Assessment (Triad CRA)[33] showed good overall agreement, although not on the specific risk stratification[34].

Whilst there are fewer RED-S studies in male athletes[2], males dancers being at risk is, consistent with findings from a self-report survey of male athletes[35] and another study of male cyclists where results from a questionnaire were linked with quantified clinical consequences of RED-S[5]. Nevertheless, the average risk score for male dancers was not as marked as in females, although fewer men responded to the questionnaire than women. Men perform a different repertoire from women, reflected in a lower percentage of men being advised to lose weight and fewer seeing low weight as a factor in being cast. Nevertheless, in common with females, the male dancers expressed a desire to be lighter and linked controlling what they ate to self-esteem.

### Limitations and further work

While there are different schools of ballet (English, Russian, French) and companies vary in their approaches, dance training follows a very similar pattern worldwide. The aim of this study was to obtain a global picture and further research may investigate dancers in a similar environment in the same company. This study was observational and cross-sectional. A forthcoming, longitudinal study will include monitoring of biometrics, performance and outcomes of educational intervention directed at behavioural change.

## Conclusions

The DEAQ indicates that dancers can display indicators of LEA, linked to high training loads, psychological drivers and external influences from initiation of dance training an early age. The DEAQ could potentially be a practical, objective screening tool to identify dancers in LEA, at risk of developing RED-S. Further prospective work is underway to combine the DEAQ with monitoring clinical biometric measures and dance performance.

## Statements

## Data Availability

Data is available upon reasonable request

## Acknowledgments

Thank you to all the dancers for their interest and participation in this study. Thanks to AusDancers Overseas for input. Thanks to One Dance UK. Thanks to teaching and research staff at Sport Science Department at Durham University.

## Contributorship

NK and AusDancers Overseas: conceptualisation of project, development of study design, involvement of dancers, drafting and revision of manuscript. GF: advanced statistical analysis drafting and revision of manuscript.

## Collaborators

AusDancers Overseas, Professor Karen Hind (Durham University)

## Data sharing

No unpublished data were used in the preparation of this manuscript

## Ethical Approval

This study was reviewed and approved by Durham University research ethics committee.

---

Supplementary file 1 Dance Energy Availability Questionnaire (DEAQ)

Supplementary file 2 Scoring system for DEAQ to assess RED-S risk

